# Artificial Intelligence-Driven Innovations in Diabetes Care and Monitoring

**DOI:** 10.1101/2025.06.02.25328795

**Authors:** Yudi Kurniawan Budi Susilo, Dewi Yuliana, Mahani Mahadi, Kapil Amgain, Shamima Abdul Rahman, Azrin Esmady Ariffin

**Author notes:** Authors’.

## Abstract

This study explores Artificial Intelligence (AI)’s transformative role in diabetes care and monitoring, focusing on innovations that optimize patient outcomes. AI, particularly machine learning and deep learning, significantly enhances early detection of complications like diabetic retinopathy and improves screening efficacy. The methodology employs a bibliometric analysis using Scopus, VOSviewer, and Publish or Perish, analyzing 235 articles from 2023-2025. Results indicate a strong interdisciplinary focus, with Computer Science and Medicine being dominant subject areas (36.9% and 12.9% respectively). Bibliographic coupling reveals robust international collaborations led by the U.S. (1558.52 link strength), UK, and China, with key influential documents by Zhu (2023c) and Annuzzi (2023). This research highlights AI’s impact on enhancing monitoring, personalized treatment, and proactive care, while acknowledging challenges in data privacy and ethical deployment. Future work should bridge technological advancements with real-world implementation to create equitable and efficient diabetes care systems.

## 1 Introduction

Artificial intelligence (AI) has emerged as a transformative force in the management and care of diabetes, enabling a range of innovations that enhance patient outcomes and optimize treatment protocols. These innovations predominantly hinge on the integration of advanced technologies, such as machine learning (ML), continuous glucose monitoring (CGM), and the Internet of Medical Things (IoMT), thereby facilitating improved self-management capabilities for patients with diabetes.

One of the most significant advancements facilitated by AI is in CGM technologies. These systems utilize AI algorithms to predict glucose levels, calibrate biosensors, and enhance closed-loop insulin delivery mechanisms, which collectively result in better glycemic control and reduced risks of hypoglycemia (Militus et al., 2024). The review by Militus et al. emphasizes how these technologies improve clinical outcomes while minimizing the administrative burdens typically associated with diabetes management, presenting a more cost-effective solution for healthcare providers (Militus et al., 2024). In conjunction with this, the use of AI-driven biosensors that monitor physiological metrics in real-time further augments diabetes care by providing timely data that can inform treatment decisions (Jin et al., 2023).

AI applications extend into predictive analytics as well, offering models capable of early detection of diabetes and its complications, such as diabetic retinopathy and nephropathy. Research indicates that AI algorithms can analyze vast datasets, including patient histories and real-time data, to identify high-risk individuals and facilitate prompt intervention (Osmanoğlu, 2023). These predictive capabilities support clinicians in making informed decisions, which is critical in managing the chronic nature of diabetes (Behera, 2021). Additionally, a comprehensive review by Alam et al. consolidates evidence from various studies indicating that ML models significantly enhance personalized management strategies, catering to individual patient responses and promoting greater adherence to treatment (Alam et al., 2024).

Moreover, AI’s role in telemedicine has been pivotal, especially in enhancing remote patient monitoring (RPM) amid the growing need for healthcare flexibility. AI-integrated telemedicine platforms enable continuous data collection from wearables and mobile health applications, facilitating ongoing patient engagement and timely modifications to treatment plans (Talati, 2023). This shift towards RPM, underscored by data from Kumar et al., showcases how the integration of AI fosters better management of diabetes by providing healthcare professionals with crucial insights into patients’ real-time health status (Kumar, 2024). Such innovations are especially crucial for managing chronic diseases like diabetes, where timely adjustments based on patient data can significantly impact health outcomes.

In specific populations, such as pregnant women with diabetes, AI technologies have devised innovative solutions that monitor and adapt to the unique challenges posed by changing insulin requirements throughout gestation (Murrin et al., 2024). The combination of AI with IoMT creates a sophisticated framework wherein healthcare providers can proactively address patient needs, underscoring the necessity of continual monitoring in this vulnerable group.

In conclusion, AI-driven innovations are reshaping diabetes care by enhancing monitoring, enabling personalized treatment protocols, and fostering proactive care environments through telemedicine and connected devices. As these technologies continue to evolve, they promise to significantly improve the management of diabetes, leading to better patient outcomes and more efficient healthcare systems overall.

## 2 Literature

Artificial Intelligence (AI) is fundamentally reshaping the landscape of diabetes care and monitoring through a variety of innovative applications that enhance diagnosis, treatment, and management of the disease. One of the most significant contributions of AI is in the area of early detection and diagnosis of diabetes-related complications. Research has demonstrated that AI algorithms can significantly improve the efficacy of screening tests, allowing for earlier and more accurate diagnoses of conditions such as diabetic retinopathy, neuropathy, and diabetic foot ulcers (Ziajor et al., 2024; Huang et al., 2022; Abràmoff et al., 2018; Chun & Kim, 2023). For instance, the FDA’s authorization of the IDx-DR system,which automatically detects more than mild diabetic retinopathy without the need for clinician interpretation, illustrates AI’s capability to provide autonomous diagnostic support in primary care settings (Abràmoff et al., 2018).

In the context of diabetes management, AI is facilitating precision medicine by enabling personalized treatment approaches based on comprehensive data analysis. Continuous Glucose Monitoring (CGM) systems, which collect real-time glucose data, when integrated with AI, can offer predictive insights that enhance glycemic control by alerting users to potential hypoglycemia or hyperglycemia (Vettoretti et al., 2020; Iftikhar et al., 2024). Such systems utilize machine learning to recognize patterns in glucose fluctuations, thereby informing patients and healthcare providers about necessary lifestyle or medication adjustments (Vettoretti et al., 2020; Jawaid & Qureshi, 2024). This ability to create data-driven insights fosters smarter insulin administration and dietary adjustments, which are crucial for effective diabetes management.

AI tools also support patient education and self-management practices, empowering individuals with diabetes to take a more active role in their care. AI-enabled platforms provide patients with tailored educational resources and feedback related to their condition, helping them manage their diabetes more effectively through personalized guidance on diet, exercise, and medication schedules (Li et al., 2020). For example, mobile applications that utilize AI can adapt recommendations based on user behavior and physiological responses, thus promoting better adherence to care protocols (Pawar & Sharma, 2019; Singla et al., 2019). This tailored approach not only helps in improving patient outcomes but also enhances overall healthcare efficiency by streamlining communication and care pathways.

Beyond direct management and education, AI is advancing research into the underlying mechanisms of diabetes and its complications. By employing deep learning and machine learning techniques, researchers are able to analyze large datasets—from genetic information to environmental factors—that contribute to diabetes progression (Vu et al., 2020; Shukur et al., 2023). This wealth of insights can lead to the identification of new biomarkers, thereby paving the way for innovative therapeutic strategies. AI’s role in observational research enhances the understanding of diabetes epidemiology, informing public health strategies aimed at prevention and risk reduction (Chemello et al., 2022; Gosak et al., 2022).

In summary, the integration of artificial intelligence into diabetes care and monitoring is not only revolutionizing how the disease is diagnosed and managed but also enhancing patient engagement and education. The potential for AI to analyze vast quantities of data to generate actionable insights exemplifies its transformative role in advancing diabetes care into a new era of precision and personalization.

## 3 Method

The methodology of this study aims to explore the role of Artificial Intelligence (AI) in driving innovations in diabetes care and monitoring, focusing on the application of AI, machine learning (ML), deep learning, and related technologies to optimize patient outcomes and improve monitoring systems. To achieve this, a combination of bibliometric analysis and data visualization tools will be employed, including VOSviewer, Harzing’s Publish or Perish, Scopus, and MS Excel.

**Figure 1.**
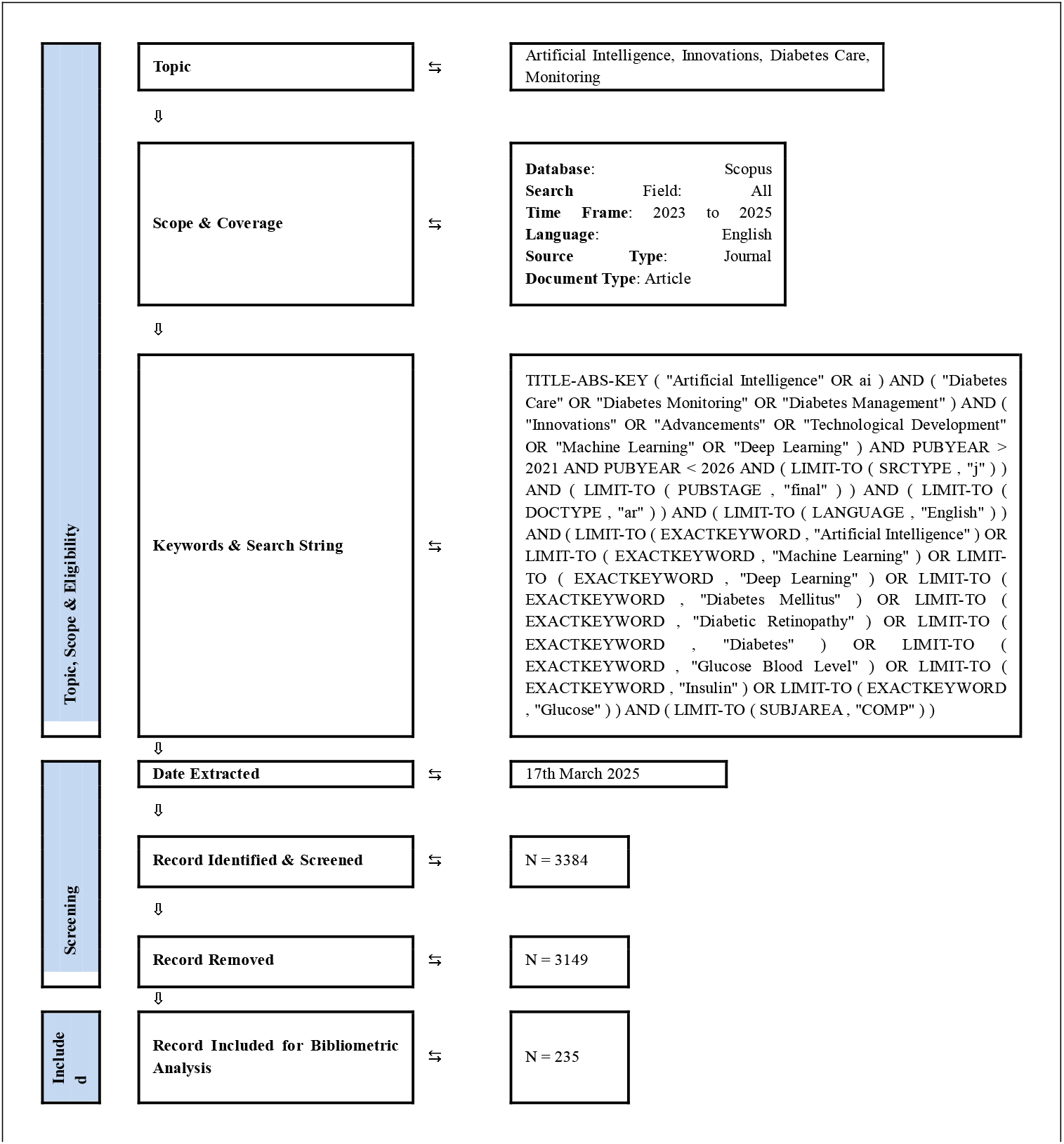
Flow diagram of the search strategy for topic Artificial Intelligence, Innovations, Diabetes Care and Monitoring.

The data collection will begin by conducting a comprehensive search in the Scopus database using a specific set of keywords such as “Artificial Intelligence”, “Diabetes Care”, “Diabetes Monitoring”, “Machine Learning”, and “Continuous Glucose Monitoring”. This search will be confined to journal articles published from 2023 to 2025, written in English. A precise search string will be used to filter out irrelevant records, ultimately yielding a dataset of approximately 3384 records. After removing duplicates and irrelevant entries, a final set of 235 articles will be selected for further analysis. The articles will focus on AI’s role in diagnosing, managing, and monitoring diabetes.

For the bibliometric analysis, the study will leverage VOSviewer, a tool for visualizing the bibliometric network. This tool will generate maps that depict relationships between authors, institutions, journals, and keywords, highlighting key trends and emerging research topics in AI-driven diabetes care. It will also help visualize how different AI technologies are interconnected within the field, making it easier to understand the research landscape.

To assess the academic impact of the identified articles, Harzing’s Publish or Perish will be used to calculate citation metrics such as the h-index, i10-index, and total citations. These metrics will provide insight into the influence of specific articles and authors in the AI and diabetes research domain. Scopus will serve as the primary source for gathering detailed bibliographic data, which will then be exported to MS Excel for further analysis.

Inclusion criteria for the data will ensure that only relevant articles are considered. The selected articles must focus on AI-driven innovations in diabetes management and monitoring, including predictive analytics, continuous glucose monitoring, AI-based diagnostics, and AI-enabled wearable devices. Studies published between 2023 and 2025 in peer-reviewed journals and written in English will be included. The research will exclude studies that fall outside these parameters.

After screening the data, the analysis will focus on several key aspects. First, keyword co-occurrence analysis will be conducted using VOSviewer to visualize the frequency and relationships between prominent keywords like “machine learning”, “diabetes”, “monitoring”, and “AI”. This will provide an understanding of the central themes in AI-driven diabetes research. Citation analysis using Publish or Perish will identify influential studies and researchers, highlighting those that have had the most significant impact on the field. Additionally, co-authorship and collaboration networks will be analyzed using VOSviewer to map the relationships between key authors and institutions, offering insights into collaborative efforts in AI and diabetes research.

Data organization and statistical analysis will be carried out in MS Excel. Descriptive statistics will summarize the key features of the dataset, such as publication trends, journal distribution, and citation counts. Trend analysis will help identify shifts in research focus, such as the growing influence of specific technologies like deep learning or the increasing use of wearable sensors in diabetes care.

Finally, the results will be presented through various visualizations, such as bar graphs illustrating publication trends over time, network maps showing keyword relationships and collaborations, and heatmaps highlighting citation patterns across journals and geographic regions. These visualizations will provide a clear and concise overview of the current state of AI-driven innovations in diabetes care and monitoring, shedding light on influential studies, emerging trends, and potential areas for future research. This methodology, combining advanced bibliometric tools and data visualization techniques, will offer a comprehensive understanding of the evolving role of AI in diabetes management.

## 4 Result

It becomes clear that the existing document distribution strongly supports the interdisciplinary nature of this topic. The largest share of documents, at 36.9% (235 documents), falls under Computer Science, which is inherently crucial for the development and application of Artificial Intelligence. Directly following in relevance, Medicine accounts for a substantial 12.9% (82 documents), underscoring the medical domain’s central role in diabetes care. Furthermore, Health Professions, at 6.6% (42 documents), highlights the practical implementation and healthcare delivery aspects of AI innovations in this field.

**Figure 2.**
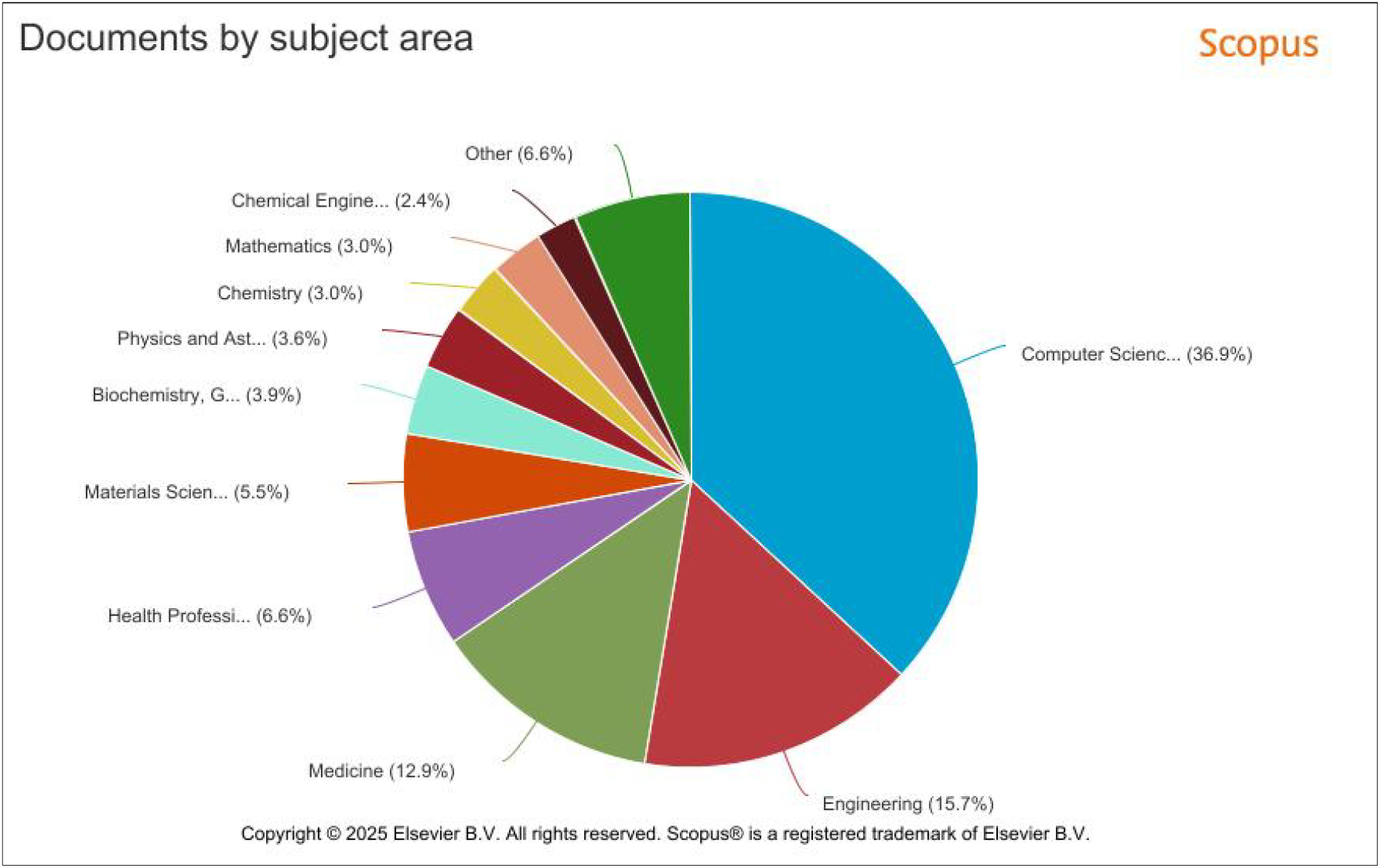
Document by subject area from Scopus repository related to topic of Artificial Intelligence, Innovations, Diabetes Care and Monitoring.

While perhaps less intuitively obvious, Engineering also holds a significant portion at 15.7% (100 documents), reflecting its contribution to the technological infrastructure, device development, and algorithmic underpinnings necessary for AI and monitoring solutions. Even Biochemistry, Genetics and Molecular Biology (3.9% / 25 documents) can play a part by informing AI models with deeper biological insights into diabetes. The remaining subject areas, while foundational to various scientific disciplines, hold less direct immediate relevance to the specific focus of AI innovations in diabetes care and monitoring.

In essence, the document distribution strongly indicates that a robust investigation into AI-driven innovations in diabetes care necessitates a deep engagement with Computer Science and Medicine, supported by contributions from Engineering and Health Professions, reflecting the multidisciplinary expertise required for such a research endeavour.

**Figure 3.**
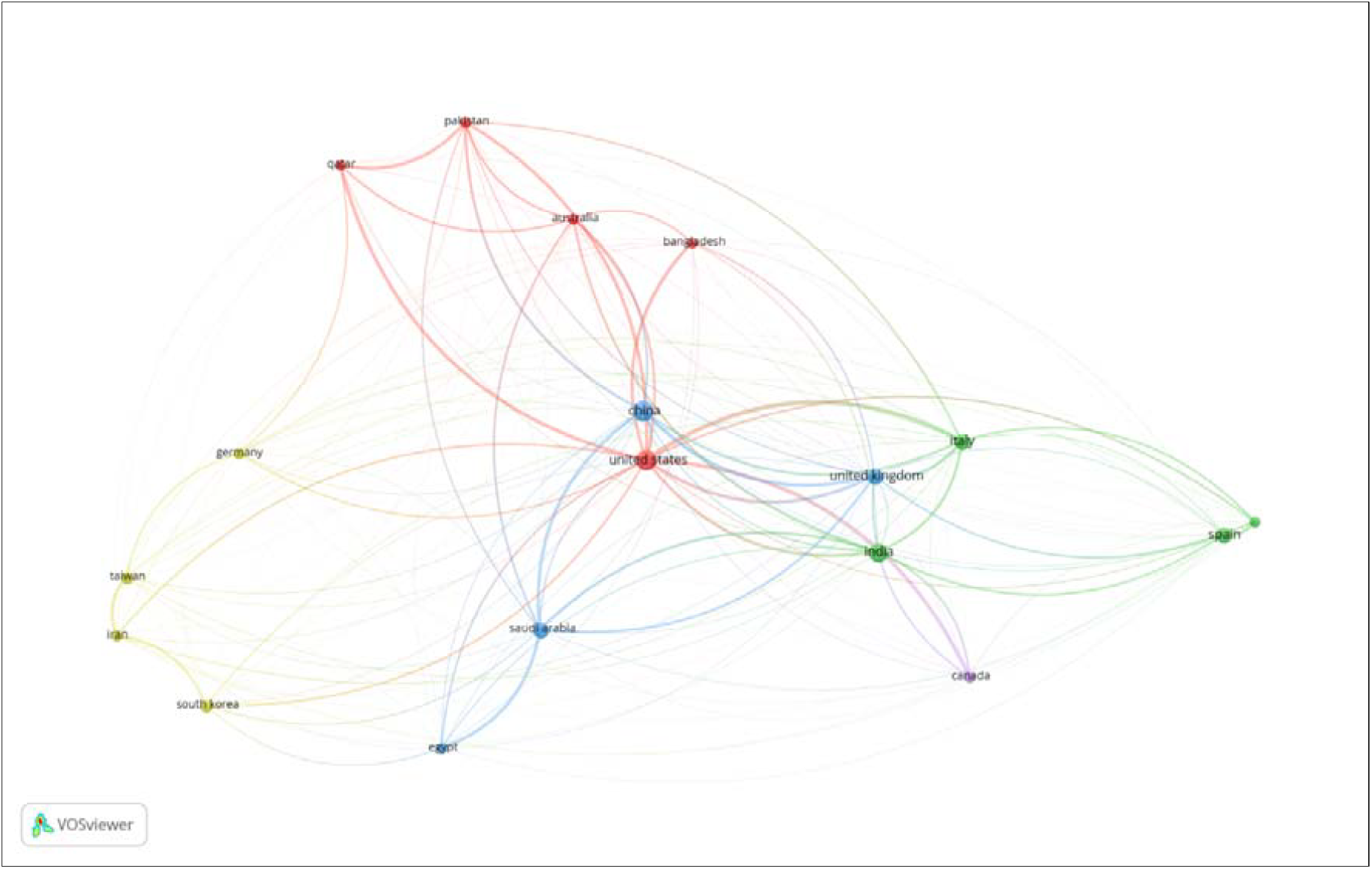
Bibliographic coupling analysis visualized the collaborative research strength among countries in the context of scholarly output.

The United States stands out significantly with the highest total link strength (1558.52), indicating its central role in research collaboration and knowledge production in AI-driven diabetes care. This dominance likely reflects the country’s strong infrastructure in both medical research and AI innovation, along with substantial funding and institutional support for interdisciplinary healthcare technologies. The United Kingdom (701.69) and Italy (684.68) follow, showcasing strong European engagement in this area. Their active participation may stem from growing public health concerns about diabetes and established research networks between medical institutions and AI labs.

China (684.48) is also a key player, nearly equal to Italy, emphasizing its rising global leadership in both artificial intelligence and biomedical research. The country’s high research output and increasing international collaborations position it as a major contributor to innovation in diabetes monitoring technologies, particularly through applications like predictive analytics and wearable AI-driven devices.

Saudi Arabia (637.17) and India (578.89) also emerge as influential contributors, likely driven by the high prevalence of diabetes in their populations, which necessitates novel healthcare solutions. These countries may be focusing on cost-effective, AI-enhanced diabetes care systems suitable for large-scale deployment in diverse healthcare settings.

Pakistan, Australia, and Qatar exhibit moderate coupling strength (ranging from 423 to 540), indicating active yet somewhat more regionally concentrated research efforts. These countries may be building strong partnerships with leading nations, particularly the U.S., U.K., and China, to co-develop or adapt AI-driven health solutions.

Meanwhile, countries like Canada, Portugal, Iran, and Egypt contribute with lower but notable link strengths, suggesting emerging or growing interest in the field. Their involvement may be catalyzed by international collaborations, academic mobility, or targeted national healthcare initiatives to tackle chronic diseases using AI.

In conclusion, the bibliographic coupling map underscores robust international collaboration network underpinning research in AI-driven diabetes care. The dominance of countries like the U.S., U.K., China, and Italy reflects not only their technological and research capabilities but also their strategic focus on using AI to enhance healthcare outcomes. For researchers and stakeholders aiming to expand or collaborate in this field, these countries represent hubs of innovation and potential partnerships for impactful work.

**Figure 4.**
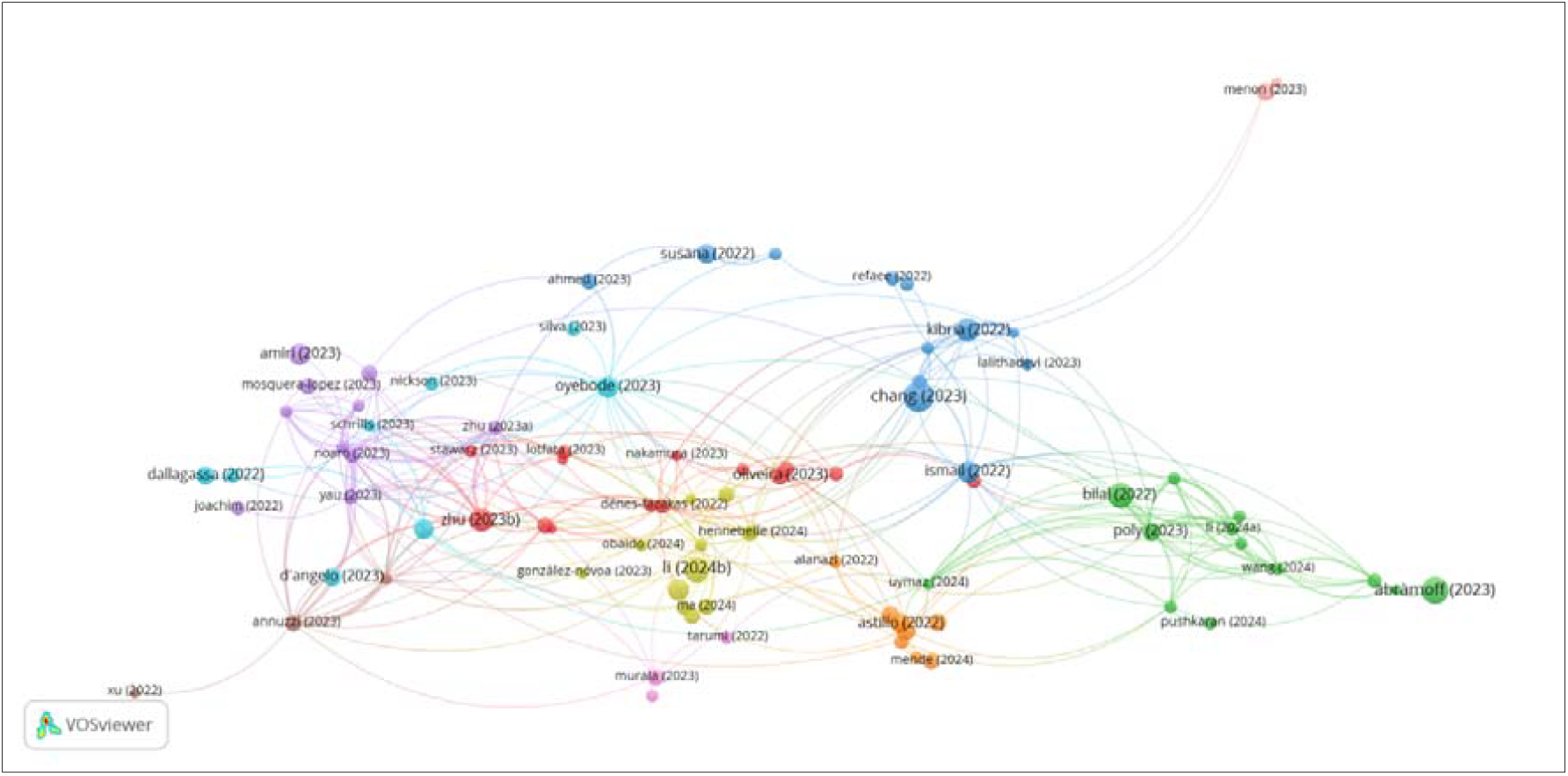
Bibliometric coupling analysis based on the VOSviewer network visualization and the ranking table of documents with the highest total link strength.

From the network diagrams, we observe clusters of documents that represent bibliographic connections, suggesting shared references or common thematic elements. Key documents with the highest total link strength include Zhu (2023c) with a strength of 57, followed by Annuzzi (2023) at 45, Annuzzi (2024) at 42, and Noaro (2023) at 40. These papers, through their strong coupling links, appear to be highly influential and interconnected with other works in the field. The dominance of these nodes in the network visualization indicates that they are central references for recent research in the area.

Given the context of AI in diabetes care, these influential papers likely explore cutting-edge technologies such as machine learning algorithms for glucose monitoring, predictive analytics for patient management, or novel sensor technologies integrated into wearable devices. The high coupling strength suggests these works are frequently co-cited or share numerous references, indicating a consolidated foundation for ongoing and future research. The presence of multiple documents by Zhu and Annuzzi points to prolific contributions that may focus on advancing computational models, real-time monitoring systems, or AI-driven patient support tools.

Moreover, peripheral nodes like Menon (2023), despite their isolated appearance, may represent specialized or emerging topics that could introduce disruptive innovations into the field. Similarly, the network’s segmentation into clusters hints at sub-themes within the broader research, such as sensor technology development, algorithm optimization, patient engagement platforms, or regulatory and ethical considerations of AI integration in diabetes care.

In summary, this bibliographic coupling analysis highlights seminal documents and thematic clusters within the research domain of AI-driven diabetes care and monitoring. The insights derived from the coupling strength and network structure suggest a vibrant and interconnected research landscape where key publications are shaping the discourse on how artificial intelligence can revolutionize diabetes management through continuous monitoring, predictive interventions, and personalized care pathways.

## 5 Discussion

The findings of this study underscore the transformative potential of Artificial Intelligence (AI) in revolutionizing diabetes care and monitoring. The interdisciplinary nature of the research, as evidenced by the dominance of Computer Science (36.9%) and Medicine (12.9%) in the document distribution, highlights the collaborative efforts required to advance AI-driven solutions. The integration of machine learning (ML), continuous glucose monitoring (CGM), and predictive analytics has significantly enhanced glycemic control, reduced hypoglycemic risks, and enabled personalized treatment protocols. For instance, AI algorithms in CGM systems, as discussed by Mililus et al. (2024), have improved real-time glucose tracking and insulin delivery, demonstrating the practical benefits of these technologies in clinical settings.

The bibliographic coupling analysis revealed strong international collaboration, with the United States, United Kingdom, and China leading in research output and innovation. This global engagement reflects the urgency of addressing diabetes as a public health challenge and the role of AI in providing scalable solutions. Key documents, such as those by Zhu (2023c) and Annuzzi (2023), have laid the groundwork for advancements in AI-driven diagnostics and wearable technologies, further validated by their high citation metrics. However, challenges remain, including the need for robust data privacy frameworks, ethical considerations in AI deployment, and the integration of these technologies into diverse healthcare systems. Future research should focus on addressing these gaps while exploring the potential of emerging technologies like deep learning and IoMT to further refine diabetes management.

## 6 Conclusion

In conclusion, this study demonstrates that AI-driven innovations are reshaping diabetes care by enhancing monitoring accuracy, enabling proactive interventions, and fostering personalized patient management. The convergence of AI with CGM, predictive analytics, and telemedicine has not only improved clinical outcomes but also empowered patients through self-management tools. The prominence of interdisciplinary research and global collaboration underscores the collective effort required to harness AI’s full potential in this field. Moving forward, stakeholders must prioritize translational research to bridge the gap between technological advancements and real-world implementation. By addressing ethical, regulatory, and accessibility challenges, AI can be leveraged to create equitable, efficient, and patient-centric diabetes care systems, ultimately improving quality of life for millions worldwide.

This study contributes to the growing body of evidence supporting AI’s role in diabetes care, while also highlighting avenues for future innovation and collaboration in this critical area of healthcare.

## Data Availability

All data produced in the present study are available upon reasonable request to the authors

## Notes

### Competing Interest Statement

The authors have declared no competing interest.

### Funding Statement

This study did not receive any funding

### Summary of Updates

author orders updated author affiliations updated

